# CTRhythm: Accurate Atrial Fibrillation Detection from Single-Lead ECG by Convolutional Neural Network and Transformer Integration

**DOI:** 10.1101/2024.10.26.24316175

**Authors:** Zhanyu Liang, Chen Yang, Zhengyang Yu, Yinmingren Fu, Bozhen Ren, Maohuan Lin, Qingjiao Li, Xuemei Liu, Yangxin Chen, Li C. Xia

**Affiliations:** Department of Statistics and Financial Mathematics, School of Mathematics, South China University of Technology, Guangzhou, China; Department of Cardiology, Sun Yat-sen Memorial Hospital of Sun, Yat-sen University, Guangzhou, China; The Eighth Affiliated Hospital, Sun Yat-sen University, Shenzhen, China; School of Physics and Optoelectronics, South China University of Technology, Guangzhou, China

**Keywords:** Atrial fibrillation, ECG classification, Convolutional neural network, Transformer encoder

## Abstract

Atrial Fibrillation (AF) is a common supraventricular arrhythmia that affects about 30 million people globally. Electrocardiogram (ECG) analysis is the primary diagnostic approach. The widespread adoption of wearable devices monitoring heart rhythm prompted the development of AF detection models for single-lead ECGs, benefitting real-time early diagnosis. Current state-of-the-art methods for AF detection are convolutional neural network (CNN) and convolutional recurrent neural network (CRNN) based models, which only focus on capturing local patterns despite heart rhythms exhibiting rich long-range dependencies. To address this limitation, we propose a novel method for single-lead ECG rhythm classification, termed CNN-Transformer Rhythm Classifier (CTRhythm), which integrates CNN with a Transformer encoder to capture local and global patterns effectively. CTRhythm achieved an overall F1 score of 0.831, outperforming the baseline deep learning models on the golden standard CINC2017 dataset. Moreover, pre-training with additional data improved the overall F1 score to 0.840. In two external validation datasets, CTRhythm showed its strong generalization capabilities. CTRhythm is freely available at https://github.com/labxscut/CTRhythm.

## I. Introduction

Atrial Fibrillation (AF) is a supraventricular tachyarrhythmia that affects about 30 million people globally [1]. Timely detection of AF is crucial for effectively managing heart conditions and preventing severe consequences [2]. Electrocardiogram (ECG) has long been a valuable diagnostic tool in identifying cardiac abnormalities, including AF. In recent years, the rapid emergence of personal cardiac monitoring devices, such as smartwatches, has led to a significant increase in single-lead ECG data [3]. Compared to 12-lead ECG data collected in hospitals, single-lead ECG data is more readily accessible through portable wearable devices. Once anomalies are detected on wearable devices, users can be prompted to seek more detailed examinations at the hospital, facilitating timely early diagnosis and treatment.

In machine learning, AF detection is commonly framed as a heart rhythm classification task [4-6]. Recently, the advent of deep neural networks significantly improved classification performance by enabling automatic feature extraction and end-to-end classification. Among deep learning methods, the convolutional neural networks (CNN) [7-13] gained prominence in AF detection primarily attributable to their efficient automatic continuous feature extraction capabilities using the convolution operator. However, CNN is inherently designed for extracting local features due to its localized receptive field of convolution kernels, making it less adept at capturing crucial contextual and global dependencies [14]. Researchers tried integrating CNN with recurrent neural networks (RNN) to address this shortcoming and proposed several CRNN-based methods [15-19]. However, those approaches require sequential traversal of the entire ECG sequence, which is inefficient and ineffective when dealing with long sequences [14].

Transformers have recently emerged as a robust model for effectively learning long-range dependencies [14]. Numerous Transformer-based models achieved remarkable success in natural language processing [20] and computer vision [21] tasks. They were also applied to ECG data for various tasks [22-24], including atrial fibrillation and flutter segmentation, heartbeat location and detection, and AF classification. Here, we propose adopting and combining Transformers with previously successful CNN models for AF detection, focusing on performance with single-lead ECG data.

In this work, we integrated CNN and Transformer to create a more comprehensive encoder and classification framework for heart rhythm classification, named CTRhythm (**Fig. 1**). CTRhythm first employed CNN to extract local features and then Transformer to encode global correspondence. The integration empowers CTRhythm to leverage the strengths of both encoders in capturing short-range and long-range rhythmic relationships. Our contributions are as follows:

**Fig. 1.**
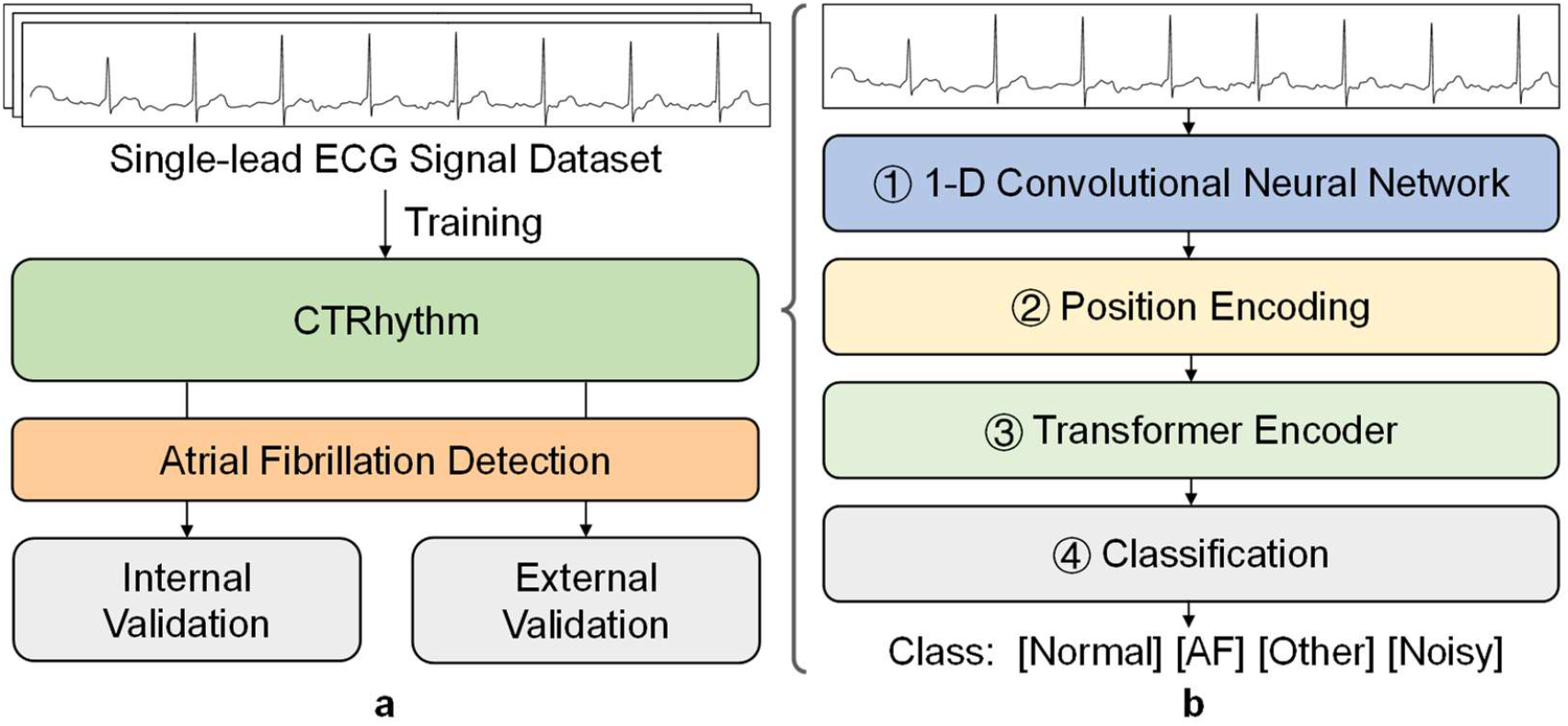
Overview of CTRhythm: **a**) the training and validation workflow; **b**) the network architecture of CTRhythm.

- We developed CTRhythm, a new rhythm classifier that effectively integrates Transformer and CNN models for AF detection.
- CTRhythm encodes local features and long-range dependencies, resulting in higher classification accuracy over baseline methods in cross-validation.
- The independent validation results in two external datasets showed that CTRhythm can generalize well.
- The ablation experiment results assessing the effect of pre-training indicated that incorporating outside data can further improve CTRhythm’s performance.

## II. Related works

### A. ECG Classification Methods

AF detection from ECG signals is commonly framed as a classification task [4-6]. In recent years, deep learning methods, such as Convolutional Neural Networks and Recurrent Neural Networks, demonstrated significantly improved performance in ECG classification tasks while eliminating the need for manual feature extraction.

CNN is the most used deep neural network in ECG classification. For example, Hannun et al. [8] developed a deep CNN to classify 12 rhythm classes in single-lead ECG. Xiong et al. [9] proposed a 16-layer 1D CNN, demonstrating improved performance by incorporating skip connections for rhythm classification.

CNNs are proven to extract local features efficiently but cannot model global contextual information because of the localized receptive field of convolution kernels. To address this limitation, researchers grafted Recurrent Neural Network (RNN) layers, such as Long Short-Term Memory (LSTM) [25], into CNNs. The resulting hybrid model, Convolutional Recurrent Neural Networks (CRNNs), proved moderately effective in classifying heart rhythm. For example, Zihlmann et al. [19] computed a one-sided spectrogram of ECG signals and applied a logarithmic transform, followed by a combination of CNN and LSTM to detect AF.

Although those hybrid models show improvement, the sequential and forgetful nature of RNN layers [14] has deteriorated performance processing long sequences, thus limiting the effective learning of global patterns. The requirement for laborious step-by-step traversal of the entire sequence also hinders RNNs from efficiently encoding long-range dependencies, significantly as the interaction of distant time points diminishes with elongated training steps.

### B. Transformer

Transformer represents a modernized approach to learning positional correspondence. It leverages a one-step attention mechanism to capture long-range dependencies and process the sequence positions in parallel. Vaswani et al. introduced the first Transformer encoder [14]. Transformers have since undergone extensive testing in natural language processing and computer vision. Their proven versatility and capability led researchers to investigate their potential applications to ECG data. For example, Yun et al. [22] developed a Transformer model for atrial fibrillation and flutter segmentation in long ECGs. Yang et al. [23] decomposed long ECGs into a series of components with clinical meanings and tokenized them. They then used a Transformer model to process these tokens for AF classification. Hu et al. [24] employed a Transformer model to simultaneously locate and detect heartbeats from ECG signals. Nonetheless, those studies have focused on applying Transformers to 12-lead hospital ECG data, and their potential value for single-lead data from wearable devices has yet to be exploited.

## III. Methods

### A. Overview

We integrated CNN and Transformer to create a novel and effective hybrid AF detection model, CTRhythm. CTRhythm consists of four modules (see Fig. 1b and Fig. 2):

1. 1D Convolutional Neural Network Module (1-D CNN): this module down-samples the input ECG sequence and aggregates the local features using CNN (Fig. 2b).
2. Position Encoding Module: this module applies positional encoding to the CNN-encoded feature sequence to encode the sequence order (Fig. 2a).
3. Transformer Encoder Module: this module performs attention-based learning of long-range dependencies on the position-CNN-encoded sequence (Fig. 2d);
4. Classification Module: this module consists of a global average pooling layer, a linear layer, and a SoftMax layer to predict scores and labels (Fig. 2a).

The details of each module are as follows:

**Fig. 2.**
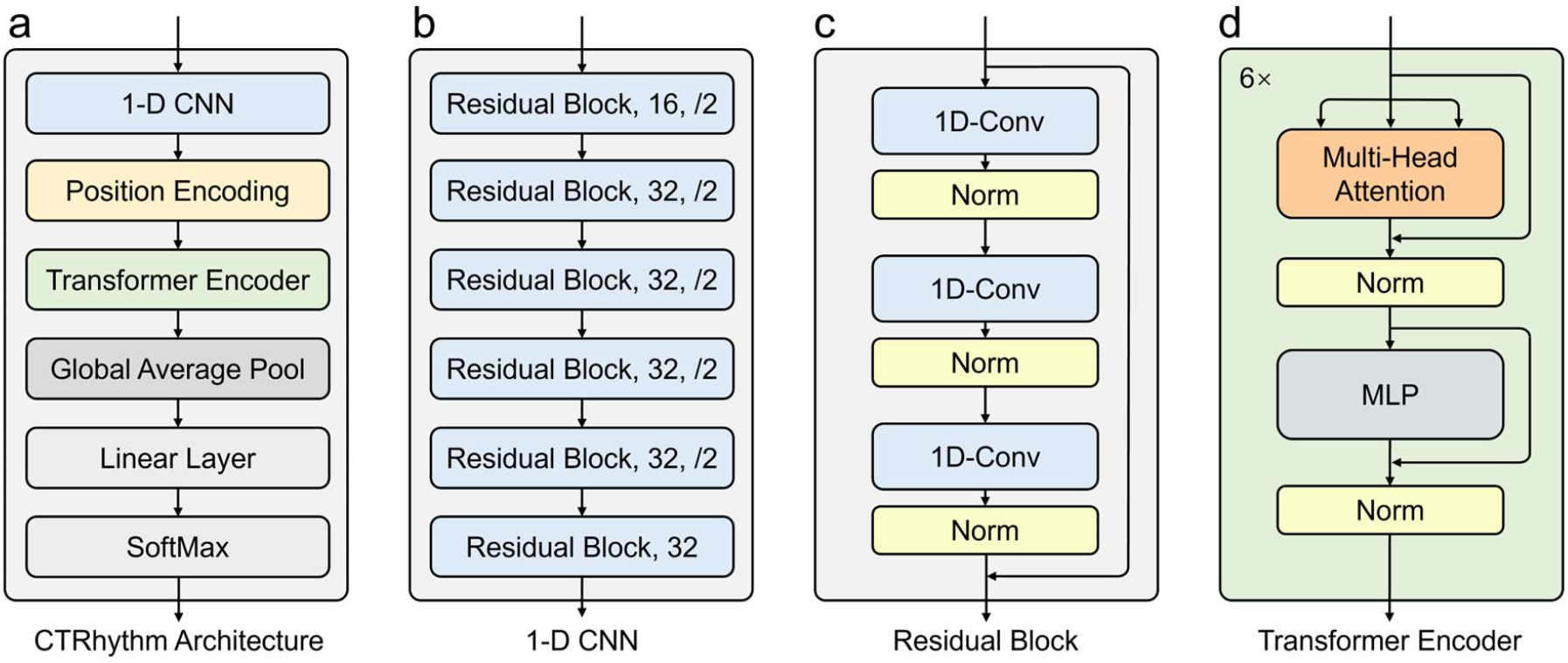
The components of CTRhythm modules: **a)** an overview of modules and data flow; **b)** the 1-D CNN module: 16 or 32 is the number of output channels, /2 is the down-sampling rate; **c)** convolution layers within one residual block of CNN; **d)** the Transformer encoder module.

### B. Convolutional Neural Network Module

Our CNN module design was inspired by ref. [26]. The critical difference is that we incorporated more down-sampling convolutional layers to effectively reduce the original sequence length. The CNN module in CTRhythm is comprised of 6 residual blocks. Each basic residual block shall consist of three 1-D convolutional layers. Batch Normalization (BN) [27] and Rectified Linear Unit (ReLU) activation layers [28] are applied after each convolutional layer (see Fig. 2c).

We employed two different residual blocks (Fig. 2b). In the first type of residual block, the convolutional kernel’s stride is set to 2, efficiently halving the sequence length. The second type of residual block maintains the shape of the input to the output sequence. Both blocks use convolutional kernels of size 3, and the number of convolutional kernels gradually increases to 32.

We stack six residual blocks to form the CNN module (Fig. 2b). Overall, the ECG sequence transforms into a feature sequence with reduced length and increased dimensions.

### C. Position Encoding

Unlike RNNs, the Transformer encoder layer neglects the positional order within the sequence [14]. Therefore, preserving the features’ order through positional encoding is crucial. Each position in the input feature sequences is linearly projected into an embedding positional latent space of dimension *d* using a learnable matrix ***W***^*E*^ ∈ ℝ^*k×d*^. In CTRhythm, *d* is set to 32, determined through experimental tuning. We used sine and cosine functions [14] as encoding bases. We denote the positional encoding of the sequence as ***E***^*PE*^:

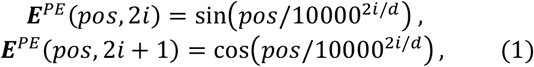

where *pos* is the position in the sequence, and *i* is the dimension. ***E***^*PE*^(*pos*, 2*i*) denotes the element in the *pos* row and 2*i* column of the matrix ***E***^*PE*^. The formula of position encoding is as follows:

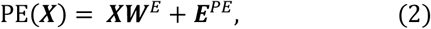

### D. Transformer Encoder

We used a stack of 6 Transformer encoder layers to form the Transformer encoder. We employed the Multi-head Self-Attention (MSA) mechanism in each layer to capture long-range dependencies. The MSA can attend to information from dimensions and positions simultaneously. In the *i*-th self-attention component, we generated the matrices ***Q***_*i*_, ***K***_*i*_ and ***V***_*i*_ as the products of ***X*** and weight matrices 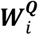, 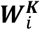 and 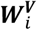, then we used a linear projection to fuse the outputs from all heads, as follows:

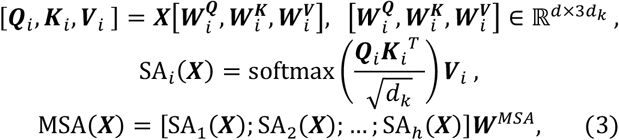

where *d*_*k*_ is the dimension of ***Q***_*i*_, ***K***_*i*_ and ***V***_*i*_, 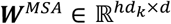, *i* = 1, 2, …, *h*, and *h* is the number of attention heads. In our model *h* is set to 8, and *d*_*k*_ is set to *d*/*h* = 4.

We used the Multi-Layer Perceptron (MLP) to facilitate additional non-linear transformations.

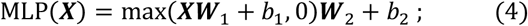

where 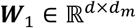, 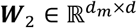, and *b*_1_, *b*_2_ are learnable parameters of two distinct linear layers. In our model, the hidden size *d*_m_ is set to 256.

After each MSA or MLP layer, Layer Normalization (LN) [29] is applied to normalize the features of each sample, thereby standardizing all input features as follows:

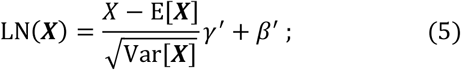

where 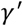 and 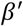 are learnable parameters adjusting the normalizing effect by this layer, and E[***X***] and 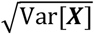 are the mean and standard deviation computed over the dimensions (along the *d* subscripts of ***X***).

We denoted the input sequence as ***X*** and the output of the *i*-th Transformer encoder layer as ***X***_*i*_, *i* = 1,2, …, 6. Let ***X***_O_ = ***X, X***_*i*_ can be iteratively computed as follows:

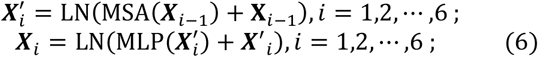

where 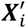 represents the intermediate output at the *i-*th layer. The last layer’s output is the Transformer encoder’s output:

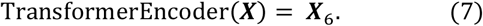

### E. Classification Module

At the end of CTRhythm, we added the classification module, which consists of a Global Average Pooling (GAP) layer [30], a linear layer, and the SoftMax function. The selection of CTRhythm’s hyperparameters is based on ref. [14], and they are adjustable based on the actual size of ECG data.

## IV. Experiment

### A. Datasets

#### The Training and Cross-Validation Dataset

The primary dataset used in our study is the PhysioNet/Computing in Cardiology Challenge 2017 (CINC2017) dataset [31]. This dataset comprises ECG recordings collected using the AliveCor, a wearable ECG device capable of recording single-lead ECG data. The training set consists of 8528 single-lead ECG recordings, encompassing four types of heart rhythms: Normal, AF (Atrial Fibrillation), Other rhythm, and Noisy. Following the experimental setup for baseline models in previous works [10, 19, 32, 33], we cross-validated CTRhythm and compared it to models using this dataset. The average F1 score from 5-fold Cross-Validation is reported as the performance metric.

#### The Two External Validation Datasets

To evaluate the generalizability of CTRhythm and other models under distribution shifts, such as those caused by data sources, we used two external validation datasets: the China Physiological Signal Challenge 2018 (CPSC2018) [34] and the Chapman datasets [35]. Those datasets were collected from hospitals and can be publicly used. We extracted all normal and AF ECG records from the two datasets. We then applied models trained on CINC2017 to those datasets.

#### The Pre-Training Dataset

To further explore the potential of CTRhythm with pre-training, we pre-trained it using the MIMIC-IV-ECG [36]. In addition to the standard internal training and validation dataset, MIMIC-IV-ECG contains about 800,000 12-lead ECG records. We only used those ECGs for self-supervised pre-training.

### B. Data Preprocessing

We down-sampled raw ECGs to 150 Hz to reduce the input size and alleviate computational burden while maintaining performance. We normalized samples by Z-score to eliminate their scale differences and shifts. The normalized data were truncated and padded to a uniform length of 60 seconds.

### C. Evaluation Metrics

To facilitate lateral comparison, we employed the standard evaluation metrics from an earlier challenge competition [31]. Considering the suboptimal quality of the Noisy class ECGs, this paper excludes them from computing the final classification performance, as done by the 2017 PhysioNet/CinC Challenge. Performance is assessed based on the average F1 score for the three categories: Normal (N), AF (A), and Other (O), denoted as F1_N_, F1_A_, and F1_0_. The overall F1 score is denoted as F1_All_, is calculated as the average of the F1 scores for all three categories. We also used accuracy to evaluate a model’s performance. In independent external validation, we used accuracy, F1_A_, F1_N_, and F1_All_ to evaluate the model performances, where F1_0_ is not applicable.

### D. Implementation Details

The experiments were conducted using PyTorch 2.1. The computational resources for training included an Intel Core i9-12900K CPU, 128 GB RAM, and an NVIDIA RTX 3090 GPU. We designed two groups of experiments to implement CTRhythm:

#### Training and Validation

All models were trained and validated using the CINC2017 dataset, with additional external validation on the CPSC2018 and the Chapman datasets. The training process consisted of 100 epochs with a batch size of 256. We employed the AdamW optimizer with a learning rate of 0.001 and a weight of decay of 0.01.

#### Pre-training, Fine-tuning, and Validation

CTRhythm was first pre-trained on the MIMIC-IV-ECG dataset. For the pre-training step, we used the MOCO v2 framework [37] with 20 epochs and a batch size of 256. The AdamW optimizer was used with a learning rate of 0.001 and a weight decay of 0.01. For the model fine-tuning on CINC2017, the learning rate was adjusted to 0.0001, while other settings remained the same as in the previous Train and Validation experiment.

## V. Results and Discussion

### A. AF detection on CINC2017

We compared CTRhythm with the most current ECG classification methods applied to the CINC2017 dataset. Table I summarises the F1 scores for all compared methods. CTRhythm achieves a state-of-the-art overall F1 score of 0.831, outperforming all the competitors.

**TABLE I.** AF detection results on the CINC2017 dataset.

| Method | Model | F1<sub>N</sub> | F1<sub>A</sub> | F1<sub>O</sub> | F1<sub>All</sub> |
| --- | --- | --- | --- | --- | --- |
| Datta et al. [32] | AdaBoost | 0.910 | 0.798 | 0.772 | 0.826 |
| Zihlmann et al. [19] | CNN | 0.878 | 0.790 | 0.701 | 0.790 |
|  | CRNN | 0.888 | 0.764 | 0.726 | 0.792 |
| Lee et al. [10] | CNN | 0.897 | 0.811 | 0.745 | 0.818 |
| Yu et al. [33] | CNN | **0.914** | 0.776 | 0.762 | 0.817 |
| ATI-CNN [38] | CNN | 0.883 | 0.717 | 0.702 | 0.767 |
| Xresnet1d101 [39] | CNN | 0.881 | 0.715 | 0.674 | 0.757 |
| MSDNN [40] | CNN | 0.900 | 0.779 | 0.746 | 0.808 |
| **CTRhythm (Ours)** | **CNN+Transformer** | **0.910** | **0.811** | **0.772** | **0.831** |

The best results are **bolded**, with gray indicating the second highest.

### B. External Validation

We used two external validation datasets to compare the generalizability of the models. Table II presents the results. Remarkably, CTRhythm outperformed all other models. Moreover, CTRhythm, trained on data from wearable devices, also performed well on data collected in hospitals, demonstrating its robustness under distribution shifts.

**TABLE II.** AF detection results under external validation.

| Dataset | Method | F1<sub>N</sub> | F1<sub>A</sub> | F1<sub>All</sub> | Accuracy |
| --- | --- | --- | --- | --- | --- |
| CPSC2018 | ATI-CNN [38] | 0.901 | 0.790 | 0.846 | 0.759 |
|  | Xresnet1d101 [39] | 0.899 | 0.764 | 0.831 | 0.749 |
|  | MSDNN [40] | 0.887 | 0.791 | 0.839 | 0.775 |
|  | **CTRhythm** | **0.968** | **0.918** | **0.943** | **0.899** |
| Chapman | ATI-CNN [38] | 0.853 | 0.797 | 0.825 | 0.728 |
|  | Xresnet1d101 [39] | 0.884 | 0.801 | 0.842 | 0.753 |
|  | MSDNN [40] | 0.892 | 0.807 | 0.849 | 0.797 |
|  | **CTRhythm** | **0.923** | **0.890** | **0.906** | **0.840** |

The best results are **bolded**, with gray indicating the second highest.

### C. Effects of Pre-training

Table III presents the results after introducing pre-training and fine-tuning steps. By incorporating ECG big data for pre-training, CTRhythm’s performance was further enhanced.

**TABLE III.** Results of pre-training.

|  | F1<sub>N</sub> | F1<sub>A</sub> | F1<sub>O</sub> | F1<sub>All</sub> | Accuracy |
| --- | --- | --- | --- | --- | --- |
| Random Init | 0.910 | 0.811 | 0.772 | 0.831 | 0.854 |
| Pre-trained | **0.916** | **0.817** | **0.788** | **0.840** | **0.862** |

### D. Visualization of Learned Features

To gain insights into our model’s learned latent ECG features, we visualized the attention values of the intermediary layers using the Attention Rollout technique [41]. The visualized attention mechanisms (Fig. 3) showed that CTRhythm learned interpretable patterns. For explainability analysis, we displayed the first sample of AF (Rec. A00004) and Normal (Rec. A00001), respectively.

**Fig. 3.**
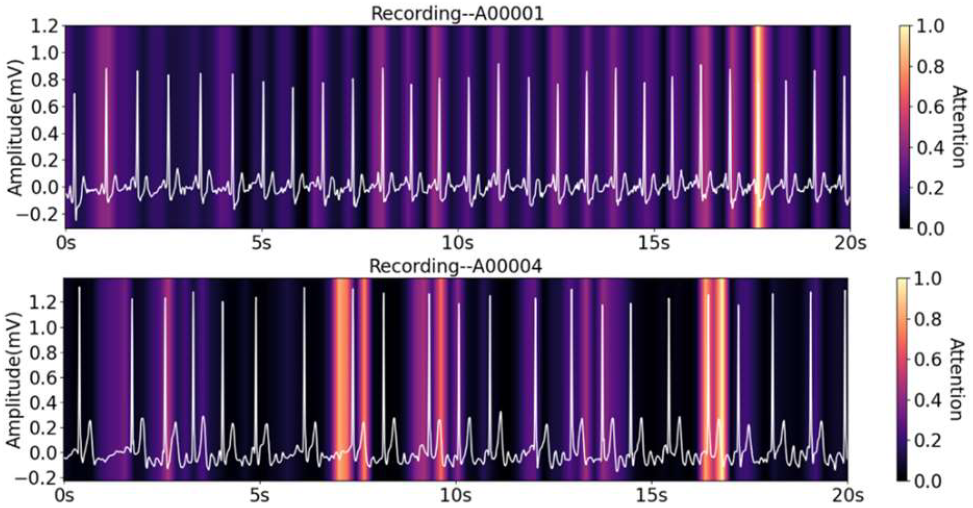
Visualization of learned features by CTRhythm. High heat areas are the locations where CTRhythm shows high attention.

In the Normal ECG, discernible P waves indicate regular atrial depolarization. The upper panel of Fig. 3 shows that CTRhythm attends evenly to these locations of the QRS complexes. In the AF ECG, atrial activation becomes chaotic, resulting in the absence of identifiable P waves and irregular RR intervals. The lower panel of Fig. 3 shows the attention heat of CTRhythm manifested in random locations.

### E. Ablation Studies

#### Ablation of Model Architecture

We performed ablation studies on the model architecture to validate the effectiveness of the Transformer encoder. The CNN model was established by removing the Transformer encoder module from CTRhythm. Subsequently, we developed a CRNN architecture by integrating the CNN model with two LSTM layers. We also created a larger CNN model by augmenting the convolutional kernels in the baseline CNN model while maintaining the parameter scale parity with CTRhythm. The cross-validated performance of these diverse architecture models is presented in Table IV, which proves the effectiveness of the Transformer encoder module.

**TABLE IV.**
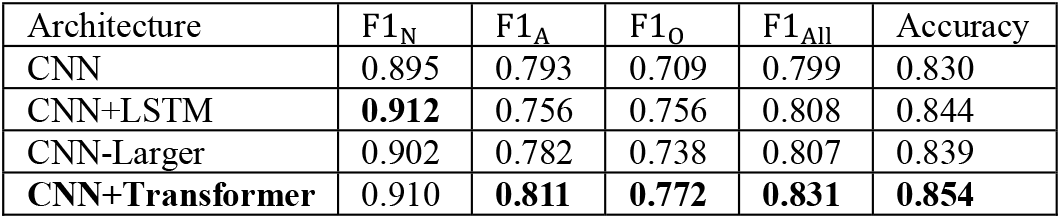
Effects of architecture choices.

#### Ablation of Model Complexity

We also conducted an ablation study on model complexity, focusing on the number of Transformer encoder layers. Table V shows the results obtained by varying the number of Transformer layers, demonstrating that CTRhythm with 6 Transformer layers achieved the highest overall F1 score (0.831) and accuracy (0.854). Interestingly, using only two Transformer layers already improved the overall F1 score of the baseline CNN by 2% (0.799 vs 0.825). This indicated that even rough integration of the Transformer module does enhance the CNN model’s ability to encode rich global rhythmic features, thus improving classification. Therefore, if the computational resource is limited, such as deploying CTRhythm to a small wearable device, a 2-layer CTRhythm is viable.

**TABLE V.** Effects of the number of Transformer layers.

| Transformer Layers | F1<sub>N</sub> | F1<sub>A</sub> | F1<sub>O</sub> | F1<sub>All</sub> | Accuracy |
| --- | --- | --- | --- | --- | --- |
| 0 (CNN) | 0.895 | 0.793 | 0.709 | 0.799 | 0.830 |
| 2 | 0.909 | 0.795 | 0.771 | 0.825 | 0.852 |
| 4 | 0.909 | 0.792 | 0.765 | 0.822 | 0.849 |
| 6 | **0.910** | **0.811** | **0.772** | **0.831** | **0.854** |
| 8 | 0.908 | 0.788 | 0.761 | 0.819 | 0.847 |

### F. Computation Requirement Analysis

We designed CTRhythm to be integrated into smartphones or wearable devices. When processing a 60-second ECG, CTRhythm requires 90 million floating-point operations and occupies ~23 MB of memory. The popular Apple A16 processor has 6 GB of memory and can perform up to 1.79 trillion floating-point operations per second. This implies that modern smartphones like iPhones can efficiently compute for AF detection in 5e-5 seconds. Additionally, if wearable devices have computation capabilities exceeding 100 million floating-point operations per second, CTRhythm could also be deployable to such devices for real-time tasks.

## VI. Conclusion

In this study, we presented a novel approach, CTRhythm, which integrates Transformer and CNN encoder models for AF detection from single-lead ECG. Unlike previous CNN or CRNN-based models, CTRhythm incorporated Transformer layers to eliminate CNN’s limits due to localized reception. By effectively encoding local and global patterns, CTRhythm outperformed all competing methods in cross-validation and showed the highest generalizability in independent external validations. Additionally, pre-training with big 12-lead ECG data further enhanced CTRhythm’s performance. Our findings highlighted the significance of effectively fusing local and global information for improved heart rhythm classification. The source code for CTRhythm is freely available at https://github.com/labxscut/CTRhythm.

## Data Availability

All data produced in the present study are available upon reasonable request to the authors
All data produced in the present work are contained in the manuscript

https://physionet.org/content/challenge-2017/1.0.0/

https://physionet.org/content/ecg-arrhythmia/1.0.0/

http://2018.icbeb.org/Challenge.html

https://physionet.org/content/mimic-iv-ecg/1.0/

## Acknowledgments

This study was funded by the Guangdong Basic and Applied Basic Research Foundation (2022A1515-011426), in part by the National Natural Science Foundation of China (61873027, 81970200, and 82271609), in part by the Guangzhou Municipal Science and Technology Project (2023B01J1011), and in part by the Shenzhen Science and Technology Program (JCYJ20190808100817047 and RCBS20200714114909234).

## Notes

### Competing Interest Statement

The authors have declared no competing interest.

### Author Declarations

The study used (or will use) ONLY openly available human data that were originally located at: https://physionet.org/content/challenge-2017/1.0.0/ https://physionet.org/content/ecg-arrhythmia/1.0.0/ http://2018.icbeb.org/Challenge.html https://physionet.org/content/mimic-iv-ecg/1.0/

